# Patient-Reported Characteristics of Pernicious Anaemia: *A first step to initiate James Lind Alliance Priority Setting Partnership Driven Research*

**DOI:** 10.1101/2024.08.30.24312837

**Authors:** Alfie Thain, Petra Visser, Kathryn Hart, Ebba Nexo, Andrew McCaddon, Luciana Hannibal, Bruce HR Wolffenbuttel, Ralph Green, Nicola Ward, Catherine Heidi Seage, Julian Owen, Katrina Burchell, Marie-Joe Dib, Kourosh R Ahmadi

## Abstract

**Objective:** Pernicious anaemia (PA) is characterised by vitamin B12 deficiency due to autoimmune-mediated loss of gastric parietal cells and intrinsic factor. The Pernicious Anaemia Society (PAS) identified 10 research priorities for PA through a James-Lind Alliance Priority Setting Partnership (JLA-PSP). This study aimed to survey PAS members to identify and characterise a cohort of patients to form a PA research repository.

**Methods:** An online survey was designed using SurveyMonkey, comprising 21 questions on diagnosis, comorbidities, family history, and management. The survey was sent to 3,482 PAS members (April-September 2022) via the PAS website and email.

**Results:** **A** total of 1,191 PAS members completed the survey. Among individuals with a probable (n=471) or suspected PA (n=500) diagnosis, 84% were UK-based, and 81% were female, with an age range of 23-93 years. Diagnosis was predominantly based on low serum B12 (50%), positive intrinsic factor (38%), and/or parietal cell autoantibodies (15%). Diagnostic delays were common, with 37% waiting ≥3 years for a diagnosis. Nearly half had one or more other autoimmune diseases. One-third reported having at least 2 and up to 7 family members with PA or other autoimmune diseases. Vitamin B12 treatment frequency varied widely, ranging from daily to 3-monthly injections.

**Conclusion:** This study highlights gaps in current diagnostic and management approaches for PA, paving the way for future work in line with the JLA-PSP research priorities. By characterising a cohort of PA patients and compiling baseline data, we provide a foundation for research to develop more effective diagnostic and management strategies.

## Introduction

Pernicious anaemia (PA) is a multifactorial, life-long disorder resulting from an incompletely understood interplay of genetic, environmental, and autoimmune factors. The disease is characterised by vitamin B_12_ deficiency caused by B_12_ malabsorption as a consequence of autoimmune atrophic gastritis leading to reduced or absent intrinsic factor (IF) production [1– 3]. Its prevalence is conservatively estimated at 0.1% in the general population, rising to 2% among those over 60 years in the UK [4,5]. Frequently coexisting with other autoimmune disorders, PA contributes to a global rise in multimorbidity [6–9].

Management for PA usually involves life-long intramuscular injections of 1 mg hydroxocobalamin every 2-3 months [10]. However, this dosing regimen lacks a robust scientific basis and many patients require more frequent injections to manage their symptoms adequately [11]. This has led to a number of patients self-managing their condition, including resorting to unofficial self-administration of intramuscular B_12_ injections [11,12]

Despite its significant health and economic impact on the patients and health-care system, PA remains largely neglected by the global research community. This is largely due to the scattered nature of its diagnosis and treatment which navigates both primary and secondary care, involving multiple medical specialities, including haematology, gastroenterology, and neurology. The establishment of the Pernicious Anaemia Society (PAS) was a direct response to these ongoing challenges [13]. As the only non-profit organisation dedicated solely to the improvement of PA diagnosis and management, PAS supports over 8,500 members and has actively supported and facilitated new research on PA. Notably, an important survey conducted by Hooper et al. in 2014 among PAS members revealed extensive diagnostic delays and dissatisfaction with treatment approaches, highlighting the urgent need for a change in PA management [14]. A more recent cross-sectional study revealed that this problem persists across several UK regions [15]. In 2020, the PAS partnered with the James Lind Alliance (JLA) to identify top research priorities through a Priority Setting Partnership (PSP), highlighting the need for more targeted research into PA’s diagnosis, treatment, and management [16]. Recent guidelines from NICE echo these priorities, emphasising the need for improved diagnostic and management protocols whilst maximising cost-effectiveness [17].

Benefitting from its unique access to a large sample of PA patients, PAS has enabled gathering of extensive patient data on diagnosis, treatment, and prognosis of PA, significantly advancing the potential for future research in this field. The current manuscript utilises the most up to date PAS database, based on data from the most recent (2022) PAS survey. We conducted exploratory analyses aimed at further highlighting gaps in knowledge with regards to diagnosis, management and to establish a new cohort of PAS members to recruit into future studies aimed at answering research questions identified by both the JLA-PSP and NICE.

## Materials and methods

### Recruitment

The survey was posted on the PAS members page and sent to all “active” PAS members (3,482 members) via email, newsletter, and website to inform them of the opportunity to participate in future research projects. An active member is considered a member who has an email associated with their membership and has consented to receiving emails from PAS. Inclusion criteria were membership of the PAS and aged 18 or above.

### Survey design

An online survey was created by the PAS and research team at the University of Surrey using Survey Monkey (SurveyMonkey Inc., San Mateo, CA, USA). It comprised twenty-one questions aimed at collecting data on demographics, mode and timing of PA diagnosis, diagnosed comorbidities, specifically autoimmune disease, family history of PA and/or other autoimmune conditions, and type/regime of treatment; the categories and details are provided in **Supplementary Table 1**.

Data were collected between April and September 2022, with all collected data transferred and stored in the PAS electronic database. Response options included yes/no, single selection, multi-tick, Likert scale or free text answers. Data from the survey was based solely on self-reported information and not confirmed by a medical examination. Each submission was checked for completeness and adherence to the survey’s requirements, with a minimum of 75% completion defined as the threshold for inclusion.

Participants were subsequently classified into the following diagnostic categories:

- Probable PA diagnosis – those diagnosed with high specificity diagnostics (a positive IFA result, gastroscopy or now obsolete Schilling test).
- Suspected PA diagnosis –a diagnosis of PA given by an HCP based on tests of lower specificity (parietal cell autoantibodies, low serum B12 levels, or elevated methylmalonic acid).
- No PA diagnosis - participants without an official PA diagnosis or those uncertain of their diagnostic status.

### Statistical analysis

Descriptive and stratified analyses were performed to investigate the associations between demographic characteristics, diagnosis timing, comorbid conditions, family medical history, and management strategies of the participants. To ensure the accuracy and relevance of the analyses, only data from participants with a probable or suspected diagnosis of PA were included.

The time from the onset of symptoms to the formal diagnosis of PA was categorised into predefined intervals to investigate the impact of diagnostic delays. Analyses also explored the relationship between familial PA status and several clinical outcomes, including treatment frequency, age at diagnosis, and the burden of autoimmune diseases. Participants were categorised based on their AID burden into no additional AIDs, one additional AID, or multiple additional AIDs.

Participants were stratified into two groups: "potentially sporadic" (reporting no other family members diagnosed with PA) and "familial" (having other family members diagnosed with PA or other AID). Linear modelling, including linear and logistic regression analysis, were used to test and quantify associations between demographic factors (age, gender), familial PA status, burden of autoimmune diseases, and frequency of B12 treatment. All statistical analyses were performed using R statistical software (version 2023.12.0+369). All analyses were conducted with a significance threshold set at p<0.05.

## Results

### Characteristics of participants

The survey link was sent to 3,482 active PAS members, of which 1,191 members (34%) responded. Among the participants, 971 (81%) reported having a diagnosis of PA, while 125 (10%) stated they did not have an official diagnosis, and 95 (8%) were uncertain.

Participants were categorised into three diagnostic groups (**Table 1**). Those with probable PA (n=471) who were diagnosed with a positive IFA (n=372, 79%), atrophic gastritis (n=98, 20%), and/or negative Schillings test (n=80, 17%) (**Figure 1**); The latter test is now obsolete and tested the patient’s ability to absorb B_12_ [18]. Patients grouped as suspected PA (n=500) presented less stringent results, including a low plasma B_12_ (n=230, 46%), a positive PCA (n=51, 10%), high MMA (n=13, 3%) or simply a diagnosis given by a HCP. Patients grouped as unlikely PA (n=220) showed none of the criteria listed for the first two groups (**Supplementary Table 3**).

**Figure 1.**
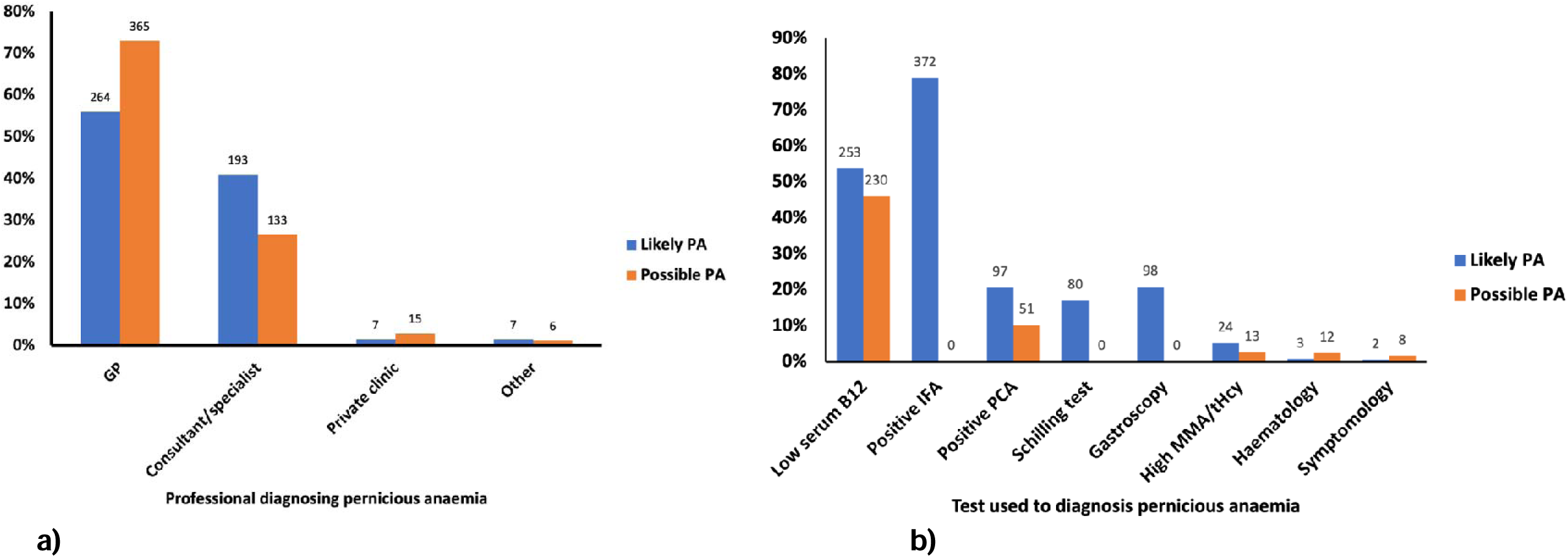
Diagnostic profiles reported by Pernicious Anaemia Society members. (a) Type of professionals diagnosing pernicious anaemia in the probable pernicious anaemia diagnosis group (n=471) and suspected pernicious anaemia diagnosis group (n=500) (b) Type of diagnostic tests used for pernicious anaemia in the probable pernicious anaemia diagnosis group (n=471) and suspected pernicious anaemia group (n=500). Haematology refers to individuals diagnosed through blood work, such as megaloblastic anaemia. Symptomology indicates that individuals are diagnosed based on clinical symptoms and/or a trial of B_12_ treatment in response to those symptoms.

**Table 1.** Diagnostic and Sex-Based Comparative Analysis of Characteristics, Diagnostics, Comorbidities, and Treatment Trends in Pernicious Anaemia Society Members (n=971).

|  | Probable PA diagnosis<sup>a</sup><br>(n=471) |  |  | Suspected PA diagnosis<sup>b</sup><br>(n=500) |  |  | Combined PA diagnoses<sup>c</sup><br>(n=971) |  |  | No PA diagnosis<sup>d</sup><br>(n=220) |  |  |
| --- | --- | --- | --- | --- | --- | --- | --- | --- | --- | --- | --- | --- |
|  | Female<br>(%) | Male<br>(%) | Total | Female<br>(%) | Male<br>(%) | Total | Female<br>(%) | Male<br>(%) | Total | Female<br>(%) | Male<br>(%) | Total |
| **Time with symptoms before diagnosis** |  |  |  |  |  |  |  |  |  |  |  |  |
| < 6 months | 32 (8) | 17 (21) | 49 (11) | 51 (14) | 9 (9) | 60 (13) | 83 (11) | 16 (9) | 109 (11) | - | - | - |
| 6 months - 1 year | 75 (20) | 16 (20) | 91 (20) | 84 (23) | 29 (30) | 114 (24) | 159 (20) | 45 (24) | 205 (21) | - | - | - |
| 1-3 years | 112 (30) | 21 (26) | 134(29) | 101 (27) | 24 (24) | 125 (27) | 213 (27) | 45 (24) | 259 (27) | - | - | - |
| 3-5 years | 46 (12) | 8 (10) | 54 (12) | 44 (12) | 12 (12) | 56 (12) | 90 (11) | 20 (11) | 110 (11) | - | - | - |
| 5 years + | 113 (30) | 20 (24) | 133(29) | 92 (25) | 24 (24) | 116 (25) | 205 (26) | 44 (24) | 249 (26) | - | - | - |
| **Vitamin B<sub>12</sub> injection frequency** |  |  |  |  |  |  |  |  |  |  |  |  |
| **Non-Standard** |  |  |  |  |  |  |  |  |  |  |  |  |
| More than weekly | 21 (6) | 6 (8) | 27 (6) | 30 (9) | 6 (9) | 37 (9) | 51 (7) | 12 (6) | 64 (7) | 23 (20) | 7 (25) | 30 (21) |
| Weekly | 31 (9) | 7 (9) | 38 (9) | 29 (9) | 6 (7) | 35 (8) | 60 (8) | 13 (7) | 73 (8) | 11 (10) | 5 (18) | 16 (11) |
| Every 2-3 weeks | 35 (10) | 11 (14) | 46 (11) | 16 (5) | 7 (8) | 23 (6) | 51 (7) | 18 (10) | 69 (7) | 8 (7) | 4 (14) | 12 (8) |
| Monthly | 76 (22) | 16 (21) | 92 (22) | 55 (17) | 17 (20) | 72 (17) | 131 (17) | 33 (18) | 164 (17) | 19 (17) | 3 (11) | 22 (15) |
| **Standard** |  |  |  |  |  |  |  |  |  |  |  |  |
| Two monthly | 82 (23) | 12 (15) | 94 (22) | 80 (24) | 15 (18) | 95 (23) | 162 (21) | 27 (15) | 189 (19) | 22 (19) | 3 (11) | 25 (17) |
| Three monthly | 105 (30) | 26 (33) | 131(31) | 121 (37) | 34 (40) | 155 (37) | 226 (29) | 60 (32) | 286 (29) | 31 (27) | 6 (21) | 38 (27) |
| **Additional Autoimmune diseases** |  |  |  |  |  |  |  |  |  |  |  |  |
| Hashimoto's Disease | 103 (27) | 6 (7) | 109(23) | 78 (20) | 16 (17) | 94 (19) | 181 (23) | 22 (12) | 203 (21) | 28 (16) | 2 (5) | 30 (14) |
| Vitiligo | 36 (9) | 7 (9) | 43 (9) | 27 (7) | 5 (5) | 32 (6) | 63 (8) | 12 (6) | 75 (8) | 9 (5) | 2 (5) | 11 (5) |
| Rheumatoid Arthritis | 28 (7) | 3 (4) | 31 (7) | 25 (6) | 14 (13) | 39 (8) | 53 (7) | 17 (9) | 70 (7) | 8 (4) | 3 (8) | 11 (5) |
| Psoriasis | 24 (6) | 5 (6) | 29 (6) | 24 (6) | 5 (5) | 29 (6) | 48 (6) | 10 (5) | 58 (6) | 14 (8) | 2 (5) | 16 (7) |
| Graves' Disease | 17 (4) | 3 (4) | 20 (4) | 18 (5) | 1 (1) | 19 (4) | 35 (4) | 4 (2) | 39 (4) | 7 (4) | 0 (0) | 7 (3) |
| Coeliac disease | 14 (4) | 2 (2) | 16 (3) | 8 (2) | 0 (0) | 8 (2) | 22 (3) | 3 (2) | 24 (2) | 2 (1) | 0 (0) | 2 (0) |
| Type 1 Diabetes | 8 (2) | 2 (2) | 10 (2) | 6 (2) | 5 (5) | 11 (2) | 14 (2) | 7 (4) | 21 (2) | 1 (1) | 1 (3) | 2 (1) |
| Any autoimmune disease* | 183 (47) | 21 (26) | 204(43) | 157 (40) | 42 (40) | 199 (40) | 340 (43) | 61 (33) | 403 (42) | 66 (37) | 9 (23) | 75 (34) |
| **Other Comorbidities** |  |  |  |  |  |  |  |  |  |  |  |  |
| Asthma | 128 (33) | 16 (20) | 144(31) | 129 (33) | 20 (19) | 149 (30) | 257 (33) | 36 (19) | 293 (30) | 59 (33) | 10 (25) | 69 (31) |
| Depression | 114 (29) | 22 (27) | 136(29) | 104 (26) | 18 (17) | 122 (24) | 218 (28) | 40 (22) | 258 (27) | 40 (22) | 12 (30) | 52 (24) |
| Arrhythmia | 53 (14) | 11 (13) | 64 (14) | 31 (8) | 12 (12) | 43 (9) | 84 (11) | 23 (12) | 107 (11) | 23 (13) | 6 (15) | 29 (13) |
| Type 2 Diabetes | 13 (3) | 13 (16) | 26 (6) | 22 (6) | 10 (10) | 32 (6) | 25 (3) | 23 (12) | 58 (6) | 9 (5) | 3 (8) | 13 (6) |
| **Micronutrient deficiencies** |  |  |  |  |  |  |  |  |  |  |  |  |
| Iron | 189 (49) | 29 (35) | 218(46) | 153 (39) | 21 (20) | 174 (35) | 342 (44) | 50 (27) | 392 (40) | 54 (30) | 9 (23) | 64 (29) |
| Folate | 101 (26) | 22 (27) | 123(26) | 93 (24) | 14 (13) | 107 (21) | 194 (25) | 36 (19) | 230 (24) | 38 (21) | 6 (15) | 45 (20) |
| Vitamin D | 125 (32) | 24 (29) | 149(32) | 101 (26) | 15 (14) | 116 (23) | 226 (29) | 39 (21) | 265 (27) | 58 (32) | 9 (23) | 67 (30) |
| **Contact for future research** | 438 (93) |  |  | 453 (91) |  |  | 891 (92) |  |  | 162 (74) |  |  |

Note: a, Probable PA Diagnosis: Includes participants diagnosed with high-specificity diagnostics, such as a positive Intrinsic Factor Antibody test, gastroscopy, or Schilling test (now obsolete).
b, Suspected PA Diagnosis: Includes participants with an official diagnosis from a healthcare professional based on tests of lower specificity, including parietal cell autoantibodies, low serum B<sub>12</sub> levels, or elevated methylmalonic acid.
c, Combined PA Diagnosis: Includes participants meeting Likely PA diagnosis and/or Possible PA diagnosis criteria.
d, No PA Diagnosis: Includes participants without an official PA diagnosis or those uncertain of their diagnostic status.
\*Other autoimmune diseases were reported including, Lichen Planus, Lichen Sclerosus, Sjogren's syndrome, Addison's Disease, Crohn's Disease, Multiple Sclerosis

Results for each of those groups are presented in (**Table 1** and **Supplementary Table 2**). No significant differences among the three groups were observed in diagnostic rates, autoimmune diseases or burden or comorbidities. However, the probable PA group had higher rates of iron deficiency (46%) compared to the suspected PA (35%, p< 0.05) and no diagnosis groups (29%, p< 0.05). The no PA diagnosis group had more participants having B_12_ injections more frequently than weekly (21%) compared to the probable PA (6%, p <0.05) and probable PA group (9%, p< 0.05). Due to the minor differences observed in the probable PA and suspected PA groups, further analysis was performed on combined data from those two groups, referred to as “combined PA” (n=971).

Of combined PA participants, 81% were female, 19% were male, and <1% reported ‘prefer not to say’. The age range of the participants was 23-93, with 86% of participants aged 50 years old or greater (**Supplementary Table 2**).

### Diagnostic characteristics of participants

Age at diagnosis ranged from 10 to 80 years for combined PA and followed a normal distribution with a mean and median of 48 years.

The majority of combined PA participants were diagnosed within primary (65%) or secondary care (34%) (**Figure 1**). Approximately 37% (n=359) of the participants reported waiting three years or longer with symptoms before receiving a diagnosis, whilst only 11% (n=109) reported waiting less than 6 months (Responses ranged from < 6 months to > 5 years) before receiving a diagnosis and starting treatment (**Table 1**). The average time since participants received their diagnosis was 14 years (SD = 10), with a range extending from less than a year to 55 years.

### Treatment Regimens

The frequency of vitamin B_12_ injections in **Table 1** illustrates the treatment frequency reported by PAS participants, ranging from daily injections to one injection every 3 months. The majority of B_12_ injections were in the form of hydroxocobalamin (70%), with other forms including cyanocobalamin (10%) and methylcobalamin (9%); Eleven per cent of participants were not sure of the injection form. Notably, 48% of participants had injections within the recommended treatment guidelines (2 or 3 monthly), while 52% reported having more frequent B_12_ injections than the guidelines. This deviation from the recommended guidelines encompassed a broad spectrum of injection frequencies, including monthly (17%), 2-3 weeks (7%), weekly (8%), and more than weekly (7%).

### Prevalence of comorbidities among participants

Among the combined PA participants, 35% reported one or more additional AID diagnoses. These ranged from a single AID diagnosis to up to 5 separate AID diagnoses. The most frequently reported AID comorbidities were Hashimoto’s disease (21%), vitiligo (8%), rheumatoid arthritis (7%), psoriasis (6%), Graves’ disease (4%), coeliac disease (3%) and type 1 diabetes (2%) (**Table 1**).

Approximately 67% of participants reported other non-AID comorbidities (**Table 1**), including asthma (30%), depression (27%), arrhythmia (11%) and type 2 diabetes (6%). In addition to B_12_ deficiency, 60% of respondents reported at least one or more other micronutrient deficiencies upon diagnosis of PA, most commonly deficiencies in iron (40%), folate (24%), and vitamin D (27%) (**Table 1**).

### Family History

Of combined PA participants, nearly one-third reported having at least one family member with PA, while 46% had at least one family member with another AID, and 19% had family members with both PA and other AIDs (**Table 2**). Generally, sisters, mothers and maternal grandmothers were the most commonly affected relatives. Additionally, 2% and 6% of participants reported having 3 or more family members with PA and AIDs, respectively (**Supplementary Table 4**).

**Table 2.** Family members with pernicious anaemia and autoimmune diseases reported by Pernicious Anaemia Society Members within the combined PA diagnosis group (n= 971).

|  | *Family members with PA* |  |  | *Family members with other AIDs* |  |  | *Family members with PA + other AIDs* |  |  |
| --- | --- | --- | --- | --- | --- | --- | --- | --- | --- |
|  | *Female (%)* | *Male (%)* | *Total (%)* | *Female (%)* | *Male (%)* | *Total (%)* | *Female (%)* | *Male (%)* | *Total (%)* |
| Mother | 95 (12) | 16 (9) | 111 (11) | 182 (23) | 21 (11) | 203 (21) | 60 (8) | 6 (3) | 66 (7) |
| Father | 44 (6) | 12 (6) | 56 (6) | 83 (11) | 17 (9) | 100 (10) | 32 (4) | 5 (3) | 37 (4) |
| Brother | 26 (3) | 2 (1) | 28 (3) | 58 (7) | 12 (6) | 70 (7) | 18 (2) | 2 (1) | 20 (2) |
| Sister | 51 (7) | 11 (6) | 62 (6) | 109 (14) | 11 (6) | 120 (12) | 35 (4) | 4 (2) | 39 (4) |
| Child | 40 (5) | 4 (2) | 44 (5) | 76 (10) | 11 (6) | 87 (9) | 28 (4) | 2 (1) | 30 (3) |
| M. Grandfather | 18 (2) | 7 (4) | 25 (3) | 27 (3) | 4 (2) | 31 (3) | 12 (2) | 6 (3) | 18 (2) |
| M. Grandmother | 61 (8) | 8 (4) | 69 (7) | 73 (9) | 4 (2) | 77 (8) | 38 (4) | 4 (2) | 42 (4) |
| P. Grandfather | 6 (1) | 3 (2) | 9 (1) | 8 (1) | 1 (1) | 9 (1) | 4 (1) | 2 (1) | 6 (1) |
| P. Grandmother | 31 (4) | 2 (1) | 33 (3) | 18 (2) | 3 (2) | 21 (2) | 16 (2) | 0 (1) | 16 (2) |
| Any family member | 264 (34) | 50 (27) | 314 (32) | 383 (49) | 63 (34) | 446 (46) | 161 (21) | 24 (13) | 185 (20) |

Note: PA, pernicious anaemia; M., Maternal; P., Paternal; Likely PA participants diagnosed with high-specificity diagnostics, such as a positive Intrinsic Factor Antibody test, gastroscopy, or Schilling test (now obsolete) and possible PA participants with an official diagnosis from a healthcare professional based on tests of lower specificity, including parietal cell autoantibodies, low serum B<sub>12</sub> levels, or elevated methylmalonic acid; A single participant can be in multiple family member categories; Any family member indicates the number of participants with at least one family member in that grouping.

### Familial versus sporadic cases of PA

Linear regression analysis was used to assess the relationship between familial PA status and the age at which individuals are diagnosed. The model estimated that individuals with familial PA are diagnosed, on average, 1.25 years earlier than those without familial PA, although this difference was not statistically significant (t= -1.35, p= 0.18).

As judged by logistic regression, participants with familial PA were less likely to receive standard treatment compared to those with sporadic PA, although the result was not statistically significant (OR = 0.86, p= 0.28). Additionally, we assessed the association between treatment type (standard vs. non-standard) and family history of autoimmune diseases (including PA) versus no family history using a chi-square test. The results showed no significant association between these variables (p > 0.05). We also tested for association between PA status (familial versus sporadic) and the burden of autoimmune diseases. Results revealed a highly significant association (x^2^= 13.97, p< 0.01), between familial PA status and autoimmune disease burden. However, logistic regression analysis adjusted for sex, age, and age at diagnosis showed no significant association between familial PA (OR 1.09, p = 0.582) and increased likelihood of autoimmune diseases.

### Sex differences in outcome data

Comparative analysis between males and females indicated no significant difference in rates of PA diagnosis, and frequency of vitamin B_12_ injections, as shown in **Table 1**. Amongst the combined diagnosis group there was a significantly higher prevalence of Hashimoto’s disease (23% vs 12%, p< 0.05), any autoimmune disease (43% vs 33%, p< 0.05), asthma (33% vs 19%, p< 0.05), and iron deficiency (44% vs 27%, p< 0.05) among females compared to males. Females were twice as likely to report having a maternal grandmother (8% vs 4%, p< 0.05) with PA as males (**Table 2**). Females demonstrated a higher prevalence of relatives with AID than males, particularly mothers (23% vs. 11%, p< 0.05), sisters (14% vs. 6%), and maternal grandmothers (9% vs. 2%).

## Discussion

In response to the publication of ten research priorities related to the cause, diagnosis, management, and treatment of PA by the JLA-PSP, our study sought to establish a PA research repository. This repository is designed to promote and facilitate research that addresses these critical questions. Our study has begun to identify trends and associations that can help form hypotheses and guide future lines of research.

### A wide range of age-of-onset is reported for PA

Generally, for diseases, the younger the age of onset, the more important the role of common environmental and/or heritable factors. Conversely, the later the age of onset, the more prominent the unique environmental factors for the individual are [19]. We used ‘age of diagnosis’ as a rough proxy for ‘age-of-onset’ in participants and observed a near-normal distribution. The findings challenged the commonly held perception that PA primarily affects older adults [4].

### Exploring the aetiology of PA

Our study participants were predominantly female (81% with female to male ratio of 4:1). This reflects the known gender imbalance in PAS membership (85% female) [20] but also the higher prevalence of autoimmune conditions among females [14,21–24].

We stratified our participants into two groups - sporadic and familial – hypothesising that different types of PA may exist. Our results suggest significant familial clustering and notably higher than rates (approximately 33% versus 20%) previously reported [25]. It is important to note that, whilst most participants identified no known familial link for their PA, this is likely offset by a high rate of under/misdiagnosis. The identification of families with multiple members with PA or other AIDs paves the way forward for future studies investigating role of rare and common genetic variants associated with susceptibility and prognosis of PA. Furthermore, sporadic and familial forms of many other multifactorial diseases exist, and the observations reported here for PA are consistent with this [26,27].

### Multimorbidity in PA Participants

Multimorbidity, defined as the presence of two or more chronic conditions, was prevalent among the participants; half reported having one or more autoimmune diseases or other comorbid conditions [28]. Additionally, although not specifically detailed in this paper, cases of complex multimorbidity involving three or more conditions affecting at least three different body systems were observed. This distinction is crucial as the management of patients with PA alone should differ significantly from those with multiple chronic conditions, such as other AIDs. Appropriate management of these coexisting conditions is essential, as mismanagement could exacerbate symptomology, leading patients to believe they require more frequent B_12_ treatment when, in fact, better management of their other conditions might be needed.

### Diagnostic groupings

The diagnostic groupings were created and assessed to determine any impact of diagnosis method on presentation. Interestingly, no major differences were found between these groups, except for higher iron deficiency in the probable PA group. This suggests that, phenotypically, the individuals present similarly regardless of diagnostic criteria. Even in the undiagnosed PA group, the presentation of the condition was similar, indicating limitations in the diagnostics used as these may be PA. However, it is important to consider that not all participants may have PA. They may have B_12_ malabsorption due to other gastrointestinal issues, such as conditions like coeliac disease and Crohn’s disease or the use of medications like proton pump inhibitors [29]. Due to these diagnostic limitations, some individuals may be categorised as requiring B_12_ injections but not truly having PA.

### What might be causing diagnostic delays in PA?

Undoubtedly, as shown by our survey, obtaining a confirmed diagnosis of PA presents a significant challenge to patients. This is partly due to diagnostic challenges, including inadequate testing protocols and lack of a ‘gold standard’ for diagnosis. There is also a lack of awareness of PA among primary and secondary care practitioners [30–32]. Our survey highlights considerable variability in the diagnostic tests administered. Most participants were diagnosed solely through assessment of serum B_12_ levels, despite recognised concerns about the sensitivity and specificity of this test [33].

Due to the diagnostic limitations, a significant proportion of individuals without official confirmation of diagnosis likely have ‘undetected’ PA, given that most need regular vitamin B_12_ injections to relieve their symptoms. The absence of the Schilling test means that individuals are currently diagnosed either through assessment of serum B_12_, B_12_ metabolic markers (MMA or tHcy), or detection of IF or PC autoantibodies, but rarely with gastroscopic confirmation [18,34].

The absence of a universally accepted ’gold standard’ for accurately diagnosing PA poses a significant obstacle to optimising patient management and presents a substantial challenge for research in this field.. The IF and gastric PC autoantibody tests suffer from compromised sensitivity and/or specificity and large platform discrepancies limiting the ability to diagnosis PA [35–37]. Several articles propose diagnostic criteria based on various tests for PA diagnosis but often lack specific or robust guidelines that describe a pathway or algorithm that clinicians can use to guide them [38,39]. Other diagnostic algorithms to guide clinicians exist, yet they incorporate tests with poor sensitivity, such as the IF autoantibody test, or lack endoscopic testing [33,40]. Furthermore, these algorithms often require the presence of objective parameters like anaemia, glossitis and a low serum B_12_, which will not be present in significant numbers of true PA cases.

Since PA is defined by the advanced and end stages of autoimmune atrophic gastritis, gastroscopy, which allows for assessing gastric mucosal changes and sampling for autoimmune atrophic gastric using the universal Sydney classification, is particularly important [41]. However, very few of our participants report being offered a gastroscopy. To support a more accurate PA diagnosis pipeline, gastroscopy should be routinely carried out with appropriate sampling techniques of the antral and corpus for histology testing. Dottori et al. propose the possible use of a serum biopsy, which could aid in determining whether a gastroscopy is warranted [39].

Timely diagnosis is critical in PA; the duration of symptoms influences symptom type, severity, and recovery [42]. Delaying B_12_ treatment for a prolonged period can result in irreversible neurological damage and permanent symptoms [38,43,44]. Vague and non-specific symptoms – due in part to autoimmune co-morbidities that present with similar symptoms - and limitations of tests, often exacerbated by lack of training and education [45], also contribute to misdiagnoses [31]. If there is a suspicion of PA but a diagnosis has not been reached, and other possible causes of the clinical presentation have been explored, it is important to trial B_12_ treatment and assess the clinical picture.

### A need for a more tailored Vitamin B_12_ treatment regimen

The varied treatment frequencies observed strongly support the need for a more tailored approach to the management of PA, which goes beyond more frequent B_12_ injections [11]. The underlying basis for these different requirements remains unknown and warrants investigation. One clear finding is the widespread dissatisfaction among respondents, with many continuing to experience B_12_ deficiency-related symptoms when following current recommended guidelines [46]. It is important to highlight that the evidence to support current guidelines is poor, based on theoretical calculations on average B_12_ excretion in the urine [47].

Biomarkers of B_12_, including serum B_12_ and MMA, are of limited value in monitoring response, or as a basis for prescribing a particular injection frequency [48]. Improved symptom monitoring and discovery of novel “response” biomarkers are, therefore, key targets for research. It will be critical to achieve tailored treatment regimens that lead to optimum symptom management and improved quality of life and prognosis for PA patients.

More than half of the participants reported receiving B_12_ injections outside of the recommended guidelines, which is likely under-reported. Anecdotal evidence suggests that recent treatment initiation, reluctance to self-inject, and financial constraints may all contribute to why more individuals are not seeking more frequent injections. A 2014 survey of PAS members reported that 13% of participants injected outside guidelines [14]. This perhaps indicates a potential shift in accessibility, knowledge and empowerment, or practitioner engagement.

### Should we recognise sub-types of PA?

We have already highlighted that, from a purely genetic perspective, there may be at least two sub-types of PA - which we named sporadic and familial. We can also begin to formulate other ways to classify PA. From our survey and previous studies, we know that approximately 50% of PA patients present with other AID comorbidities, including vitiligo and autoimmune thyroid disease (Grave’s & Hashimoto’s’), whilst in 50% of individuals, PA remains their only diagnosis.

The mechanisms of association between PA and other AID are unknown. The life course of PA allows stratification of the disease into those factors occurring prior to diagnosis (e.g., including *Helicobacter pylori* infection) or those associated as a consequence and life-course of the disease itself (i.e. iron, vitamin D or folate deficiency). Finally, we can also clearly describe sub-types of the disease based on treatment (type, mode) requirements. Follow-up studies using the proposed repository described here will aid identification of other factors that could be used to stratify PA into different sub-types, contributing towards a new ‘precision medicine’ approach to future PA diagnosis and management.

### Proposed objectives of the PAS Research Repository

The primary objective is to support research aligned with JLA-PSP priorities in PA and recent NICE guidelines[49]. Additionally, we aim to enhance participant recruitment through strategies developed with healthcare institutions and patient advocacy groups, focusing on integrating datasets from diverse sources into current and future studies. We also plan to facilitate medical records review and harmonise metadata from established biobanks, working with healthcare providers to securely access detailed medical histories, diagnoses, and treatment information. Finally, we will establish protocols for the collection, processing, and storage of biological samples (blood, urine, DNA/RNA), ensuring informed consent and addressing ethical considerations in collaboration with healthcare facilities.

New research to improve our understanding of the range of symptoms experienced by individuals with PA and their varying responses to treatment is urgently needed. The development of objective biomarkers to assess treatment response and to understand the basis of certain individuals require more frequent B_12_ injections will address 7 out of the 10 questions of the JLA-PSP. The exploration of multi-omics-based technologies, including metabolomics and proteomics, the utility of wearables (smart-watches), and neurological tracking markers as tools to identify novel markers that objectively assess symptomology and response to treatment in PA, should be considered as priority aims [50,51].

### Guidelines for researchers wanting to use the repository to tackle JLA questions

To facilitate research inquiries and collaborations utilising the PA Research Repository, we are in the process of forming an executive committee, comprised of representatives from the PAS committee, Club-12 committee, researchers from the University of Surrey, and patients. Researchers interested in accessing the repository for studies aligned with the JLA research priorities or other related topics are encouraged to initiate contact through the corresponding author of this manuscript in the first instance.

### Strengths and limitations

The PAS research repository offers a unique resource with the potential to facilitate research, advance knowledge, and improve patient quality of life. The screening survey has provided valuable initial data and identified a willing cohort for future studies. However, there are several limitations to consider.

Firstly, the data is primarily self-reported, introducing potential bias. Participants were exclusively recruited from PAS, which may not fully represent the broader PA population. This selective recruitment could indicate a higher motivation to manage their condition, potentially reflecting a more severe disease profile, dissatisfaction, or higher socioeconomic and educational status [52]. While the participant group is geographically representative of the UK (participants are predominantly UK-based), negative experiences may bias the dataset. However, this cohort includes individuals reporting high satisfaction with their treatment regimen despite their involvement and willingness to participate in research. We currently lack granular data on medication usage for comorbidities, which could provide valuable insights into overall disease management. More detailed diagnostic test results are also needed, particularly for individuals without a formal diagnosis. However, obtaining this data may be challenging without access to medical records. Future research will prioritise individuals with probable PA diagnoses. We aim to further investigate and characterise these individuals as uncertain, contributing to a better understanding of PA diagnosis, as results from those without a diagnosis showed that they had undergone insufficient testing.

## Conclusion

Our survey of PAS members has established the first ever PA research repository of over 1,000 patient participants and paved the way to address key questions outlined by the JLA-PSP and 2024 NICE Guidelines. An overview of the baseline data has given initial insight into the complexities of research into PA, from varied age-of-onset and familial clustering to diagnostic challenges and treatment variability. The potential complexity of PA, shown by the existence of distinct forms with varied aetiologies and manifestations, highlights the need for personalised screening and clinical care tailored to the specific form of PA or its presentation. This would improve diagnosis and ensure that management plans are optimally aligned with each patient’s unique health profile, thereby advancing towards a more precision-driven model in PA diagnosis and management.

### CRediT author statement

**Alfie Thain:** Conceptualization, Methodology, Formal analysis, Writing - Original Draft. **Petra Visser:** Conceptualization, Methodology, Resources. **Kathryn Hart:** Supervision, Writing - Original Draft. **Ebba Nexo:** Conceptualization, Writing - Review & Editing. **Andrew McCaddon:** Writing - Review & Editing. **Luciana Hannibal:** Writing - Review & Editing. **Bruce HR Wolffenbuttel:** Writing - Review & Editing. **Ralph Green:** Writing - Review & Editing. **Nicola Ward:** Writing - Review & Editing. **Catherine Heidi Seage:** Writing - Review & Editing. **Julian Owen:** Writing - Review & Editing. **Katrina Burchell:** Writing - Review & Editing, Resources. **Marie-Joe Dib:** Writing - Review & Editing. **Kourosh R Ahmadi:** Conceptualization, Formal analysis, Methodology, Writing - Original Draft, Supervision.

## Supporting information

the categories and details are provided in Supplementary Table 1.

## Data Availability

All data produced in the present study are available upon reasonable request to the authors

