## Supplementary material for "Patient-Reported Characteristics of Pernicious Anaemia: *A first step to initiate James Lind Alliance Priority Setting Partnership Driven Research*": the categories and details are provided in Supplementary Table 1.

| **Category** | **Details** |
| --- | --- |
| General | Sex, birth year, country of residence, and ethnicity |
| Diagnosis | Medically confirmed PA diagnosis by HCP, diagnostic tests used, outcomes of diagnostic tests, duration of symptoms before diagnosis, age at diagnosis. |
| Management and Vitamin B12 treatment | Who supplies the treatment, type of vitamin B_12_ treatment, form of vitamin B_12_, frequency of treatment, length of time receiving treatment, Satisfaction with treatment |
| Family medical history | Family with PA, family with autoimmune conditions |
| Comorbidities | Autoimmune diseases, other diseases, nutritional deficiencies |

***Supplementary* Table 1:** Survey Data Categories and Collected Information

***Supplementary* Table 2.** Comparative Analysis of Demographics in Pernicious Anaemia Society Members (n=971)

|  | **Probable PA diagnosis^a^**  **(n=471)** | **Suspected PA diagnosis^b^**  **(n=500)** | **Combined PA diagnoses^c^**  **(n=971)** | | | **No PA diagnosis^d^**  **(n=220)** |
| --- | --- | --- | --- | --- | --- | --- |
| ***Sex*** |  |  |  |  |  |  |
| Female | 388 (82) | 395 (79) |  | 783 (81) |  | 179 (81) |
| Male | 82 (18) | 104 (21) |  | 186 (19) |  | 40 (18) |
| **Age** |  |  |  |  |  |  |
| 20-29 | 6 (1) | 10 (2) |  | 16 (2) |  | 3 (1) |
| 30-39 | 11 (2) | 14 (3) |  | 25 (3) |  | 10 (5) |
| 40-49 | 54 (11) | 40 (8) |  | 94 (10) |  | 26 (12) |
| 50-59 | 126 (27) | 109 (22) |  | 235 (24) |  | 52 (24) |
| 60-69 | 152 (32) | 147 (29) |  | 299 (31) |  | 64 (29) |
| 70-79 | 101 (21) | 135 (27) |  | 236 (24) |  | 50 (23) |
| 80-89 | 21 (4) | 43 (9) |  | 65 (7) |  | 12 (5) |
| 90+ | 0 (0) | 2 (<1) |  | 2 (<1) |  | 3 (1) |
| ***Country of residence*** |  |  |  |  |  |  |
| United Kingdom | 399 (88) | 417 (92) |  | 816 (84) |  | 158 (90) |
| USA | 27 (6) | 16 (4) |  | 43 (4) |  | 6 (3) |
| Ireland | 7 (2) | 3 (1) |  | 10 (1) |  | 0 (0) |
| Canada | 3 (1) | 6 (1) |  | 9 (1) |  | 1 (1) |
| Australia | 9 (2) | 6 (1) |  | 15 (2) |  | 3 (2) |
| Other | 7 (2) | 6 (1) |  | 13 (2) |  | 8 (5) |

Note: a, Probable PA Diagnosis: Includes participants diagnosed with high-specificity diagnostics, such as a positive Intrinsic Factor Antibody test, gastroscopy, or Schilling test (now obsolete).

b, Suspected PA Diagnosis: Includes participants with an official diagnosis from a healthcare professional based on tests of lower specificity, including parietal cell autoantibodies, low serum B12 levels, or elevated methylmalonic acid.

c, Combined PA diagnosis: Combined PA Diagnosis: Includes participants meeting Likely PA diagnosis and/or Possible PA diagnosis criteria.

d, No PA Diagnosis: Includes participants without an official PA diagnosis or those uncertain of their diagnostic status.

***Supplementary* Table 3**. Diagnostic tests reported by participants classified as ‘no diagnosis’ of Pernicious Anaemia (n= 125).

| Tests | N (%) |
| --- | --- |
| Serum B12 | 84 (67) |
| Intrinsic factor antibody | 45 (36) |
| Parietal cell antibody | 15 (12) |
| Schilling test | 1 (1) |
| Endoscopy | 14 (11) |
| MMA | 13 (10) |
| Homocysteine | 10 (8) |

***Supplementary* Table 4.** Total number of family members with pernicious anaemia and other autoimmune diseases in Pernicious Anaemia Society members in the combined PA diagnoses group (n=971)

|  | ***Family members with PA*** | | | ***Family members with other AIDs*** | | |
| --- | --- | --- | --- | --- | --- | --- |
| **N Family members** | ***Female (%)*** | ***Male (%)*** | ***Total (%)*** | ***Female (%)*** | ***Male (%)*** | ***Total (%)*** |
| 0 | 519 (66) | 136 (73) | 657 (68) | 400 (51) | 123 (66) | 525 (54) |
| 1 | 181 (23) | 37 (20) | 218 (22) | 221 (28) | 45 (24) | 266 (27) |
| 2 | 64 (8) | 12 (6) | 76 (8) | 105 (13) | 15 (8) | 120 (12) |
| 3 or more | 19 (2) | 1 (2) | 20 (2) | 57 (6) | 3 (2) | 60 (6) |

Note: PA, pernicious anaemia; AID, Autoimmune disease; Combined PA Diagnosis: Includes

Likely PA participants diagnosed with high-specificity diagnostics, such as a positive Intrinsic Factor Antibody

test, gastroscopy, or Schilling test (now obsolete) and possible PA participants with an official diagnosis from a healthcare professional based on tests of lower specificity, including parietal cell autoantibodies, low serum B_12_ levels, or elevated methylmalonic acid.
